# Application of landmark analysis and piecewise Cox regression to identify features associated with prognosis: a national retrospective cohort study of New Zealand women

**DOI:** 10.1101/2025.03.18.25324230

**Authors:** B Woodhouse, W Laux, A Trevarton, A Lasham, N Knowlton

## Abstract

**Background:** Breast cancer prognosis changes over time in complex ways depending on individual risk factors. This study aimed to analyze how breast cancer outcomes in New Zealand women change over time and identify features associated with breast cancer specific survival and locoregional recurrence across different receptor subtypes.

**Methods:** A retrospective cohort study was conducted using data from Te Rēhita Mate Ūtaetae (Breast Cancer Foundation National Register) on 21,574 women diagnosed with invasive breast cancer between 2000-2019. We applied k-medians survival clustering, landmark analysis, and piecewise Cox regression to identify time-specific risk patterns and prognostic features.

**Results:** Survival improved significantly for women diagnosed more recently. Triple-negative breast cancer had the poorest 5-year breast cancer specific survival but demonstrated better outcomes for women surviving beyond this period. In contrast, ER+/HER2-tumors, associated with favorable short-term outcomes, showed the highest risk of late recurrence and breast cancer mortality beyond 10 years. Younger age at diagnosis (≤44 years) was associated with increased recurrence risks, especially for ER-/HER2+ tumors. Radiation therapy reduced early LRR across subtypes. Tumor grade was inversely associated with late recurrence, while stage 2 disease in ER+ tumors markedly elevated late recurrence odds compared to stage 1.

**Conclusions:** This study demonstrates the dynamic nature of breast cancer prognosis, with key findings emphasizing the time-dependent shifts in risk across receptor subtypes. These findings underscore the importance of personalized, receptor-specific follow-up strategies, including extended monitoring for subgroups at heightened long-term risk.

## 1. Introduction

Breast cancer is the most common cancer in women worldwide with the highest incidence and mortality rate (Amato et al., 2023; Newman, 2023). Aotearoa New Zealand (NZ) has one of the highest rates of breast cancer in the world and demonstrates a 16% excess mortality rate (Arnold et al., 2022; Aye et al., 2023). There are approximately 3,500 new breast cancer diagnoses in NZ each year (*Te Whatu Ora - Cancer Web Tool*, n.d.) with 1 in 9 women diagnosed over their lifetime (New Zealand. Te Aho o te Kahu, 2020), and approximately 650 breast cancer deaths each year (*Te Whatu Ora - Cancer Web Tool*, n.d.). Promisingly, outcomes for women diagnosed with breast cancer in NZ have improved over the past 20 years, with 91% of women living beyond 5 years (Gautier et al., 2022). Therefore, there is considerable interest in determining what features are associated with recurrence and survival beyond 5 years (Gautier et al., 2022). Through the analysis of long-term prognostic factors, a deeper understanding can be gained of how risk patterns evolve over time, providing insights into recurrence and survival that are less influenced by biases from early disease progression or treatment-related mortality.

Prognosis in breast cancer is not static; it evolves over time due to the interplay of tumor biology, treatment, and patient-specific factors. Risk is further stratified by molecular subtypes, based on expression levels of hormone receptors. Overexpression is defined as increased expression above normal levels of cell receptors such as estrogen receptor (ER), progesterone receptor (PR) and human epidermal growth factor receptor 2 (HER2). An increase in receptor levels may increase the cancer’s sensitivity to hormone and targeted therapies. Breast cancers are initially classified by their hormone receptor status as ER+PR+, ER+PR-, ER-PR+ or ER-PR-, where + means that a proportion of tumor cell nuclei above a defined threshold are positive (i.e. staining is visible) (Leake et al., 2000; *RCPA - Breast Biopsy ER and PR Receptor*, *s*n.d.). Notably, the definition for ER or PR positivity changed in 2010 from 10% to 1% cells staining (Hammond et al., 2010).

On a population level, breast cancer has been previously studied in NZ (Brown et al., 2017; Curtis et al., 2005; Gautier et al., 2022; Gurney et al., 2020; Lao et al., 2019; Lawrenson et al., 2016, 2017, 2019; Lawrenson, Lao, Campbell, Harvey, Seneviratne, et al., 2018; McKenzie et al., 2011; Meharry et al., 2021; Seneviratne, Campbell, Scott, Shirley, & Lawrenson, 2015; Seneviratne, Campbell, Scott, Shirley, Peni, et al., 2015; Seneviratne et al., 2016; Seneviratne, Lawrenson, et al., 2015; Tin Tin et al., 2018) demonstrating associations between breast cancer specific survival and patient features (e.g. ethnicity, age, detection method) or disease features (e.g. stage, pathological subtype). Breast cancer survival disparities between ethnicities in NZ are well reported (Curtis et al., 2005; Gautier et al., 2022; Gurney et al., 2020; Seneviratne, Campbell, Scott, Shirley, Peni, et al., 2015; Seneviratne, Lawrenson, et al., 2015; Tin Tin et al., 2018), with Māori (HR 1.76; 95% CI: 1.51– 2.04) and Pacific women (HR 1.97; 95% CI: 1.67–2.32) having higher risk of mortality from breast cancer (Tin Tin et al., 2018), particularly in younger wāhine (female) Māori (Curtis et al., 2005). Asian women have previously been shown to have better breast cancer specific survival than European women (HR 0.64; 95% CI: 0.49-0.82) (Gautier et al., 2022; Lao et al., 2019). These studies suggested breast screening, stage at diagnosis, deprivation level, and type of treatment as contributors to these ethnic disparities (Curtis et al., 2005; Gurney et al., 2020; Seneviratne, Campbell, Scott, Shirley, Peni, et al., 2015; Tin Tin et al., 2018). In contrast, they suggested that biological tumor features (e.g. molecular subtype) played a less significant role in explaining the ethnic differences in outcomes (Seneviratne, Lawrenson, et al., 2015). No significant survival differences were found between ethnicities when only analyzing women with screen-detected breast cancers (Lawrenson et al., 2019; Seneviratne, Campbell, Scott, Shirley, & Lawrenson, 2015), despite Māori having significantly worse 10-year survival compared to European women with non-screen detected cancer (Seneviratne, Campbell, Scott, Shirley, & Lawrenson, 2015), which highlights the importance of screening. When considering location and access to treatment, Lawrenson et al. showed that while overall there was no difference between breast cancer mortality between women who live rurally compared to urban, worse breast outcomes were found for Māori women living rurally compared to urban (Lawrenson et al., 2016). Women with triple negative breast cancers had the worst survival of all receptor subtypes (79% 10-year survival) (Gautier et al., 2022; Lawrenson, Lao, Campbell, Harvey, Seneviratne, et al., 2018), and women with ER+/HER2-breast cancers had the best survival (92% 10-year survival) (Gautier et al., 2022). Breast cancer survival differences have also been shown across age groups, with women diagnosed 45-69 years having better 10-year survival (89%; 95% CI 89-90%) compared to women diagnosed younger (82%) or older (80%) (Gautier et al., 2022).

In 2022, an authoritative report on breast cancer in NZ was released (Gautier et al., 2022), presenting a broad analysis on cohort and tumor characteristics, treatment, and outcome, for people diagnosed with breast cancer between 2003-2019. Of relevance to this study were the hazard ratios (HRs) which showed a continuous improvement over time for women diagnosed across this 16-year period, which was observed for all ages, ethnicities, and disease stages. However, the breadth of this report did not allow for an examination of the long-term risk factors for women with breast cancer and how they are likely to shift after and/or during prolonged systemic treatment.

To address this lack of insight into long-term risk factors and changes over time, we have applied both statistical and machine learning methodologies capable of managing and interpreting time-related changes in data. These include k-medians survival clustering, landmark analysis, piecewise Cox regression, and logistic regression. These allow for a comprehensive analysis of prognostic features that are distinct from those typically captured by standard Cox regression models. For example, k-medians survival clustering is an unsupervised machine learning method that can be used to identify groups with similar survival (Villanueva et al., 2019a). Landmark analysis can be used to examine how features associated with risks for women with breast cancer might change over time, by removing all participants who were censored or died prior to a designated landmark time (Bansal & Heagerty, 2019; Morgan, 2019). Internationally, there are numerous examples of landmark analysis in breast cancer (Alafchi et al., 2019; Hotton et al., 2023; Parast & Cai, 2013; Xu et al., 2022), which have shown, for example, that there are more deaths after 5 years from HER2-breast cancers than HER2+ breast cancers with the same ER/PR status, with no difference detectable in the absence of a landmark. Piecewise Cox regression can also be applied to utilize multiple landmarks to segment the follow-up time into contiguous, mutually exclusive intervals, and outputs different estimates of risk for each interval of time (Quantin et al., 1999; Wong et al., 2017). This method incorporates the strengths of landmark analysis, while also retaining some survival dynamics over time. Traditional logistic regression can also be used to evaluate the relationship between a binary variable (such as recurrence) and a set of covariates although its assumptions can mask time related effects.

In this study, we used these advanced techniques to analyze a national breast cancer cohort from NZ, exploring outcomes for women with invasive breast cancer over time. Survival clustering was used and validated with adjusted Kaplan-Meier curves to assess survival differences by diagnosis year. This is the first NZ-based study to evaluate the effects of clinicopathological features—such as age, ethnicity, detection method, receptor subtype, and tumor grade—on locoregional recurrence (LRR) and breast cancer-specific survival (BCSS) at 5- and 10-year intervals using landmark analysis and piecewise Cox regression. Additionally, multivariable logistic regression was employed to examine the relationship between clinicopathological features and both early (≤5 years) and late (≥10 years) LRR, shedding light on actionable factors that could inform long-term patient monitoring and tailored interventions.

## 2. Material and methods

### 2.1 Data collection

Data for this study was sourced from Te Rēhita Mate Ūtaetae (Breast Cancer Foundation National Register, subsequently referred to as Te Rēhita), which includes comprehensive clinicopathological and survival data for breast cancer patients. Deidentified data, including demographic, treatment, and outcome variables, was released following review by Te Rēhita’s Clinical Advisory Group. Ethical approval was obtained from the Auckland Health Research Ethics Committee (AHREC) (Ref. AH2800) before the commencement of research.

### 2.2 Study population

This study analyzed women with stage 1-4 breast cancers (representing invasive breast cancers, including both ductal and lobular carcinomas) (n=26,463) in NZ diagnosed between 2000-2019 in four main regions, Auckland (Northern), Waikato (Midland), Wellington (Central) and Christchurch (Southern), covering at least 63% (Gautier et al., 2022) of the population (Figure 1).

**Figure 1.**
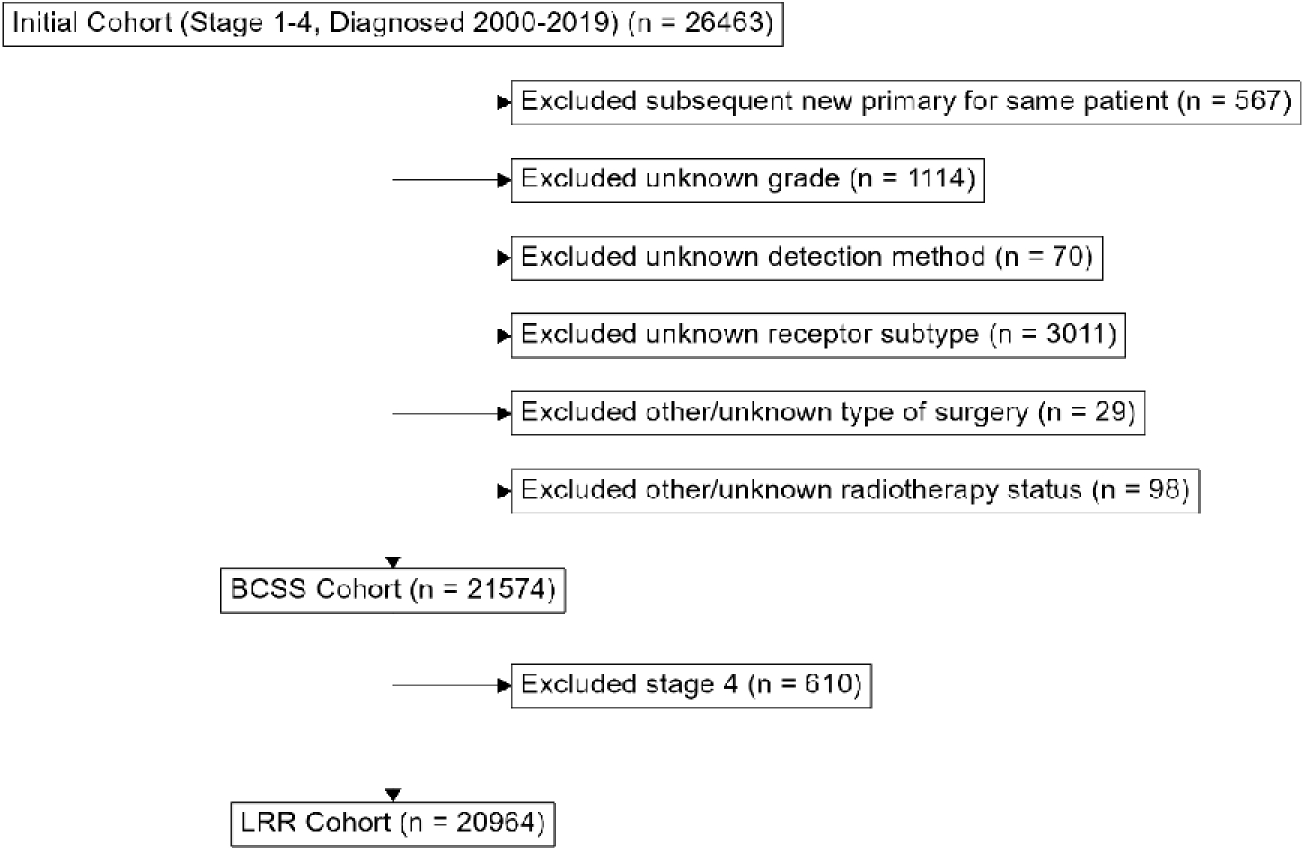
Flow diagram describing the study cohort and showing the number of women excluded in each category. This study included women with invasive breast cancers only (stage 1-4) diagnosed 2000-2019 to allow for sufficient follow-up time for survival analysis. After exclusions for missing data or new primary cancers, 21,574 women were included in analyses of breast cancer-specific survival (BCSS). Those women with stage 4 disease were excluded prior to analyses of LRR.

The timeframe was selected to ensure a minimum of 3 years of follow-up and minimize potential biases associated with the 2020 expansion of Te Rēhita. Patient follow-up was deemed current as of March 2, 2023. BCSS was calculated as the time from diagnosis to either death due to breast cancer (event) or censoring at death from other causes or the end of the study period. Median follow-up for BCSS was 8 years, including both those who experienced the event (death from breast cancer) and those who were censored. Median follow-up was 8.5 years for those patients who were alive (censored) at the end of the study period. LRR-free survival was defined as the interval between date of diagnosis and the appearance of either local or regional recurrence. The median length of time women in this cohort (LRR Cohort, no women with stage 4 disease) were recurrence free was 7.8 years.

### 2.3 Statistical analysis

Statistical analyses were conducted to evaluate BCSS and LRR-free survival. K-medians survival clustering was performed using the *survclustcurves* function from the *clustcurves* R package (Villanueva et al., 2019b) to identify groups of diagnosis years with similar survival patterns. This analysis utilized 200 bootstrap samples to ensure robustness and was limited to a maximum of 10 years of BCSS. The algorithm tested three potential cluster values (k = 2, 3, 4) and selected the optimal number of clusters. Clustering was conducted for all diagnosis years and separately for subsets defined by receptor subtypes. Adjusted Kaplan-Meier curves were used to assess survival clustering of diagnosis year groups, plotted using the ggadjustedcurves function from the survminer R package (Kassambara et al., 2021).

Landmark analysis was employed to compare survival and risk profiles for women who survived beyond predefined time points (5 or 10 years) with those of the full cohort. Adjusted curves were plotted for both the full cohort, and those who survived 5+ years BCSS, using adjustedCurves::adjustedsurv() and Direct Standardization method (Chang et al., 1982; Makuch, 1982) to adjust for age group, grade, stage and receptor subtype. For women who survived past a given landmark, survival probabilities for the pre-landmark period were set to 1.0, reflecting their survival status. However, the exclusion of patients with less follow-up time than the required landmark led to increased uncertainty in early-timepoint estimates and reduced statistical power for those intervals.

Piecewise Cox regression was used to analyze time-dependent changes in risk by dividing follow-up time into contiguous intervals (<5 years, 5–10 years, and >10 years). Separate Cox regression models were fitted for each interval to estimate HRs for BCSS and LRR-free survival (Quantin et al., 1999; Wong et al., 2017). This method retains survival dynamics over time while incorporating the strengths of landmark analysis. For LRR-free survival, follow-up time was similarly divided into <5 years, 5–10 years, and >10 years. To account for regional variation, particularly the introduction of Southern and Central regions in 2009 and 2010, respectively, region was included as a stratum variable in all multivariable models.

To ensure consistency, breast cancer cases were staged using the 7th edition of the American Joint Committee on Cancer (AJCC 7) TNM staging criteria. This decision was made because earlier cases in the cohort lacked sufficient information for AJCC 8 staging. All statistical analyses were conducted using R version 4.3.1.

## 3. Results

### 3.1 Study cohort

A summary of the clinical characteristics of study participants is presented in Table 1. The median age at diagnosis was 58 years and ranged from 20 to 98 years, with 68.5% (14,779/21,574) of women being between 45-69 years at diagnosis. The majority of women were European (n=15,917/21,574, 73.8%), with Māori comprising 9.7% (2,094/21,574) of the cohort, followed by Asian (1,883/21,574, 8.7%) and Pacific women (1,291/21,574, 6.0%). The proportion of Asian women diagnosed increased over time (p<0.05; Chi-sq test for trend), while the proportion of diagnoses for other ethnicities appeared more stable. Most invasive breast cancers were detected symptomatically (11,847/21,574, 54.9%), rather than by breast screening mammogram. The most common tumor receptor subtype was ER+/HER2-(16,177/21,574, 75.0%) which appeared to increase over time, although the definition of an ER or PR positive tumor was changed in 2010 from 10% to 1% of cells staining positive, which may have contributed to this increase (Hammond et al., 2010). The majority of breast cancer cases in this cohort were grade 2 (10,217/21,574, 47.4%) and stage 1 (12,370/21,574, 57.3%), with 1,900 (5.8%) women diagnosed with advanced breast cancer (stage 4). Analysis of the types of surgery performed showed that the proportion of women receiving breast conserving surgery compared to Mastectomy increased over time (p<0.05; Chi-sq test for trend).

**Table 1.** Summary table describing characteristics of women with invasive breast cancer in Te Rēhita Mate Ūtaetae (n=21,574), with covariates in rows and diagnosis year groups in columns. Diagnosis year is condensed into groups of three years, with the final group representing two years (2018-19). Proportion within each year group is shown in brackets for categorical variables, and standard deviation (SD) within brackets for the continuous variable (age at diagnosis). Women with breast cancer detected following symptoms (as opposed to breast screening) were defined as having detection method “Not Screened”. Breast cancer cases were staged according to the 7th edition of American Joint Committee on Cancer TNM staging criteria. The subgroups are ordered for consistency in subsequent analyses, which logically have the reference groups first, denoted with a ⍰.

|  | 2000-2002<br>(n=492) | 2003-2005<br>(n=1,593) | 2006-2008<br>(n=2,559) | 2009-2011<br>(n=3,826) | 2012-2014<br>(n=4,717) | 2015-2017<br>(n=4,989) | 2018-2019<br>(n=3,398) | Overall<br>(n=21,574) |
| --- | --- | --- | --- | --- | --- | --- | --- | --- |
| **Age at Diagnosis (years)** |  |  |  |  |  |  |  |  |
| Mean (SD) | 54.6 (14.1) | 55.8 (13.0) | 57.1 (13.1) | 57.0 (12.5) | 57.7 (12.4) | 59.0 (12.6) | 58.8 (12.4) | 57.8 (12.7) |
| Median [Min, Max] | 54.0 [20.0, 92.0] | 55.0 [25.0, 97.0] | 56.0 [21.0, 94.0] | 56.0 [22.0, 95.0] | 57.0 [21.0, 94.0] | 59.0 [20.0, 97.0] | 58.0 [23.0, 98.0] | 57.0 [20.0, 98.0] |
| **Age at Diagnosis group (years)** |  |  |  |  |  |  |  |  |
| 45-69 ☐ | 275 (55.9%) | 1,032 (64.8%) | 1,688 (66.0%) | 2,663 (69.6%) | 3,307 (70.1%) | 3,453 (69.2%) | 2,361 (69.5%) | 14,779 (68.5%) |
| 20-44 | 136 (27.6%) | 318 (20.0%) | 432 (16.9%) | 577 (15.1%) | 630 (13.4%) | 569 (11.4%) | 378 (11.1%) | 3,040 (14.1%) |
| 70-98 | 81 (16.5%) | 243 (15.3%) | 439 (17.2%) | 586 (15.3%) | 780 (16.5%) | 967 (19.4%) | 659 (19.4%) | 3,755 (17.4%) |
| **Ethnicity** |  |  |  |  |  |  |  |  |
| European ☐ | 399 (81.1%) | 1,213 (76.1%) | 1,874 (73.2%) | 2,878 (75.2%) | 3,544 (75.1%) | 3,618 (72.5%) | 2,391 (70.4%) | 15,917 (73.8%) |
| Māori | 40 (8.1%) | 144 (9.0%) | 258 (10.1%) | 381 (10.0%) | 433 (9.2%) | 477 (9.6%) | 361 (10.6%) | 2,094 (9.7%) |
| Pacific Peoples | 17 (3.5%) | 80 (5.0%) | 159 (6.2%) | 227 (5.9%) | 276 (5.9%) | 320 (6.4%) | 212 (6.2%) | 1,291 (6.0%) |
| Asian | 20 (4.1%) | 116 (7.3%) | 207 (8.1%) | 281 (7.3%) | 392 (8.3%) | 490 (9.8%) | 377 (11.1%) | 1,883 (8.7%) |
| Other/Unknown | 16 (3.3%) | 40 (2.5%) | 61 (2.4%) | 59 (1.5%) | 72 (1.5%) | 84 (1.7%) | 57 (1.7%) | 389 (1.8%) |
| **Detection Method** |  |  |  |  |  |  |  |  |
| Screened ☐ | 140 (28.5%) | 543 (34.1%) | 1,006 (39.3%) | 1,804 (47.2%) | 2,204 (46.7%) | 2,393 (48.0%) | 1,637 (48.2%) | 9,727 (45.1%) |
| Not Screened | 352 (71.5%) | 1,050 (65.9%) | 1,553 (60.7%) | 2,022 (52.8%) | 2,513 (53.3%) | 2,596 (52.0%) | 1,761 (51.8%) | 11,847 (54.9%) |
| **Region (domicile at diagnosis)** |  |  |  |  |  |  |  |  |
| Northern ☐ | 340 (69.1%) | 1,246 (78.2%) | 1,982 (77.5%) | 2,124 (55.5%) | 2,375 (50.3%) | 2,501 (50.1%) | 1,671 (49.2%) | 12,239 (56.7%) |
| Central | 0 (0%) | 0 (0%) | 0 (0%) | 521 (13.6%) | 808 (17.1%) | 873 (17.5%) | 578 (17.0%) | 2,780 (12.9%) |
| Midland | 152 (30.9%) | 347 (21.8%) | 577 (22.5%) | 549 (14.3%) | 655 (13.9%) | 704 (14.1%) | 535 (15.7%) | 3,519 (16.3%) |
| Southern | 0 (0%) | 0 (0%) | 0 (0%) | 632 (16.5%) | 879 (18.6%) | 911 (18.3%) | 614 (18.1%) | 3,036 (14.1%) |
| **Receptor Subtype** |  |  |  |  |  |  |  |  |
| ER+/HER2-<sup>®</sup> | 282 (57.3%) | 1,025 (64.3%) | 1,807 (70.6%) | 2,871 (75.0%) | 3,645 (77.3%) | 3,790 (76.0%) | 2,757 (81.1%) | 16,177 (75.0%) |
| ER+/HER2+ | 79 (16.1%) | 173 (10.9%) | 233 (9.1%) | 351 (9.2%) | 450 (9.5%) | 567 (11.4%) | 329 (9.7%) | 2,182 (10.1%) |
| ER-/HER2+ | 48 (9.8%) | 139 (8.7%) | 183 (7.2%) | 194 (5.1%) | 215 (4.6%) | 192 (3.8%) | 116 (3.4%) | 1,087 (5.0%) |
| Triple Negative | 83 (16.9%) | 256 (16.1%) | 336 (13.1%) | 410 (10.7%) | 407 (8.6%) | 440 (8.8%) | 196 (5.8%) | 2,128 (9.9%) |
| **Tumor Grade** |  |  |  |  |  |  |  |  |
| 1<sup>®</sup> | 77 (15.7%) | 242 (15.2%) | 650 (25.4%) | 962 (25.1%) | 1,078 (22.9%) | 1,026 (20.6%) | 775 (22.8%) | 4,810 (22.3%) |
| 2 | 240 (48.8%) | 763 (47.9%) | 1,137 (44.4%) | 1,705 (44.6%) | 2,219 (47.0%) | 2,465 (49.4%) | 1,688 (49.7%) | 10,217 (47.4%) |
| 3 | 175 (35.6%) | 588 (36.9%) | 772 (30.2%) | 1,159 (30.3%) | 1,420 (30.1%) | 1,498 (30.0%) | 935 (27.5%) | 6,547 (30.3%) |
| **Disease Stage (AJCC 7)** |  |  |  |  |  |  |  |  |
| 1<sup>®</sup> | 254 (51.6%) | 818 (51.3%) | 1,421 (55.5%) | 2,237 (58.5%) | 2,663 (56.5%) | 2,977 (59.7%) | 2,000 (58.9%) | 12,370 (57.3%) |
| 2 | 190 (38.6%) | 611 (38.4%) | 895 (35.0%) | 1,245 (32.5%) | 1,615 (34.2%) | 1,621 (32.5%) | 1,159 (34.1%) | 7,336 (34.0%) |
| 3 | 31 (6.3%) | 99 (6.2%) | 143 (5.6%) | 207 (5.4%) | 298 (6.3%) | 290 (5.8%) | 190 (5.6%) | 1,258 (5.8%) |
| 4 | 17 (3.5%) | 65 (4.1%) | 100 (3.9%) | 137 (3.6%) | 141 (3.0%) | 101 (2.0%) | 49 (1.4%) | 610 (2.8%) |
| **Adjuvant Radiotherapy** |  |  |  |  |  |  |  |  |
| No<sup>®</sup> | 185 (37.6%) | 542 (34.0%) | 902 (35.2%) | 1,292 (33.8%) | 1,636 (34.7%) | 1,816 (36.4%) | 1,131 (33.3%) | 7,504 (34.8%) |
| Yes | 307 (62.4%) | 1,051 (66.0%) | 1,657 (64.8%) | 2,534 (66.2%) | 3,081 (65.3%) | 3,173 (63.6%) | 2,267 (66.7%) | 14,070 (65.2%) |
| **Surgery Type** |  |  |  |  |  |  |  |  |
| Breast Conserving Surgery<sup>®</sup> | 203 (41.3%) | 738 (46.3%) | 1,207 (47.2%) | 1,846 (48.2%) | 2,335 (49.5%) | 2,664 (53.4%) | 1,914 (56.3%) | 10,907 (50.6%) |
| Mastectomy | 289 (58.7%) | 855 (53.7%) | 1,352 (52.8%) | 1,980 (51.8%) | 2,382 (50.5%) | 2,325 (46.6%) | 1,484 (43.7%) | 10,667 (49.4%) |

### 3.2 Analysis of breast cancer-specific survival and locoregional recurrence-free survival over time

To study breast cancer outcomes by year of diagnosis, standard Cox proportional hazard models adjusted for patient age, disease stage and tumor grade were used to generate Kaplan-Meier (KM) curves to examine any differences by diagnosis year. This showed that BCSS and LRR-free survival tended to improve with each successive year (Figure 2A, 2B). K-medians survival clustering was then used to group diagnosis years with similar survival profiles. This showed that women diagnosed after 2009 had improved BCSS compared to women diagnosed 2005-2008, and women diagnosed 2005-2008 had better BCSS to those diagnosed prior to 2005 (Figure 2C). These distinct groups were less apparent for LRR events (Figure 2D). Analysis of the demographic features across these diagnosis-year clusters showed a decreasing trend in the proportion of younger women diagnosed over time, while the proportion of Asian women diagnosed increased (Supplementary Table 1). Analysis of clinicopathological features across the diagnosis-year clusters showed an increase in screen-detected breast cancers, and a rise in the proportion of ER+/HER2-subtypes and Stage 1 diagnoses over time (Supplementary Table 1).

**Figure 2:**
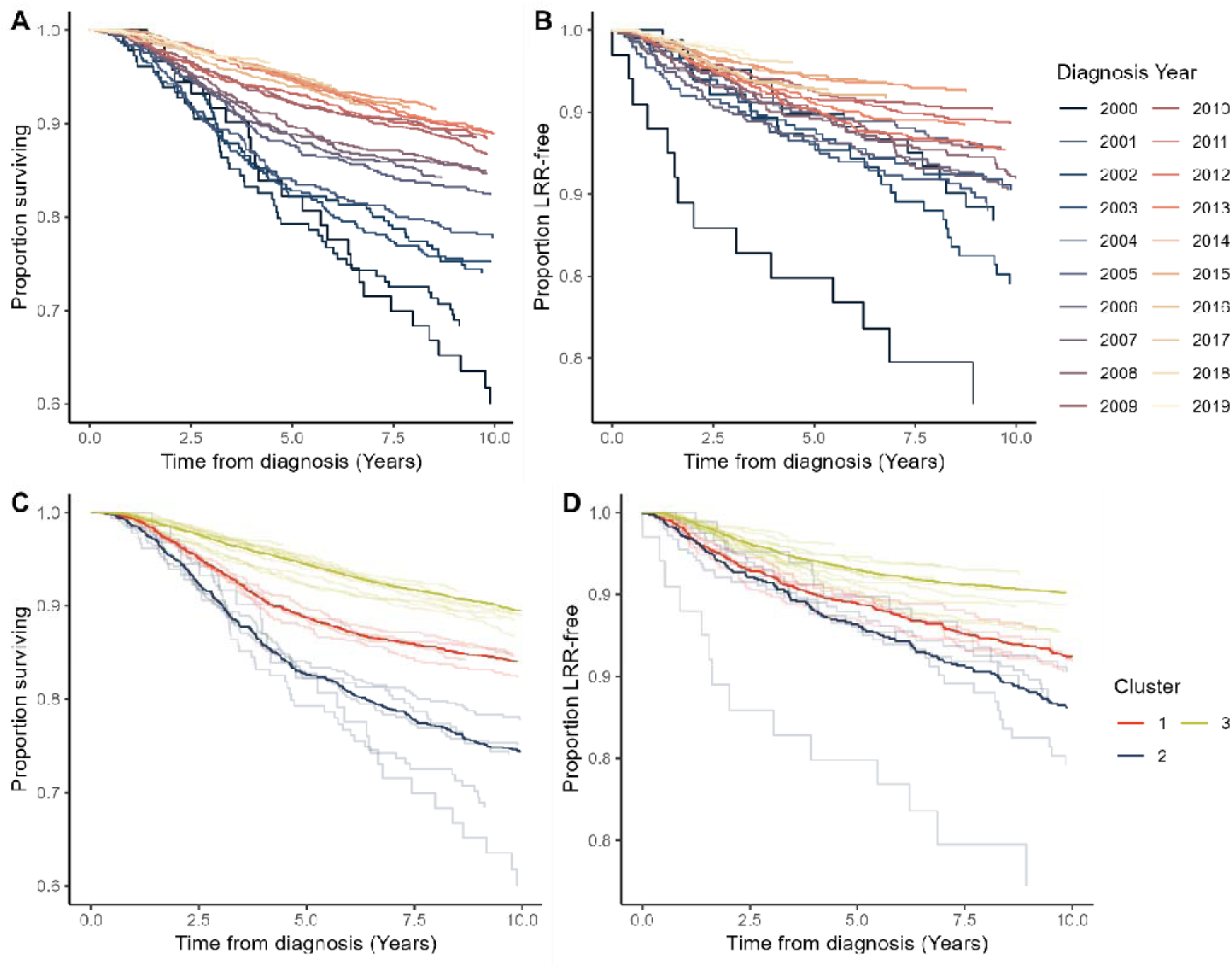
Kaplan-Meier curves by year of diagnosis and diagnosis year cluster, where clusters represent year groups with similar median survival. Both BCSS A) and LRR-free survival B) in general improved with increasing diagnosis year. K-medians survival clustering on BCSS revealed breakpoints between 2004-05 and 2008-09. Kaplan-Meier curves for BCSS (C) and LRR-free survival (D) were colored by these same breakpoints, and adjusted for patient age, disease stage, tumor grade and receptor subtype.

### 3.3 Piecewise Cox regression

To identify the changes in risk factors over time, piecewise Cox regression was used to evaluate the risk of breast cancer-specific mortality and LRR within critical intervals: 0-5 years, 5-10 years and over 10 years after diagnosis of invasive disease. For women diagnosed at 70 years or older, the overall risk of dying from breast cancer was elevated (HR 1.38) compared to women diagnosed between 45-69 years. Yet this risk profile changed over time with similar risk within the first 5 years (HR 0.98), reduced risk between 5-10 years (HR 0.62), and slightly elevated risk after 10 years (HR 1.18, Table 2). For women diagnosed before age 45 years, their overall risk of dying from breast cancer was similar to women aged 45-69 years, but when analyzed by time, younger women had a significantly greater risk of dying within 5 years of diagnosis (HR 1.24) compared to older women (Table 2).

**Table 2.**
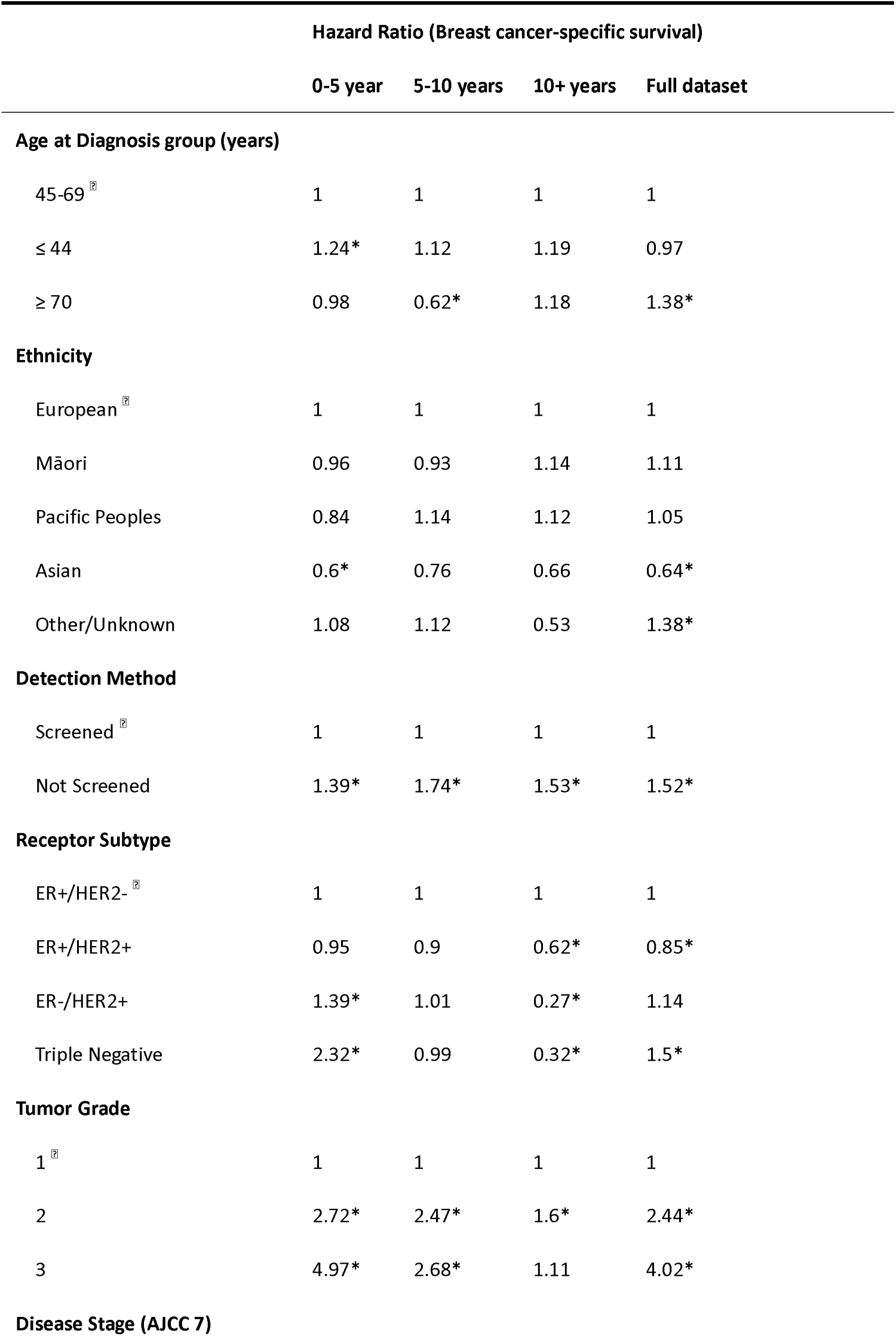
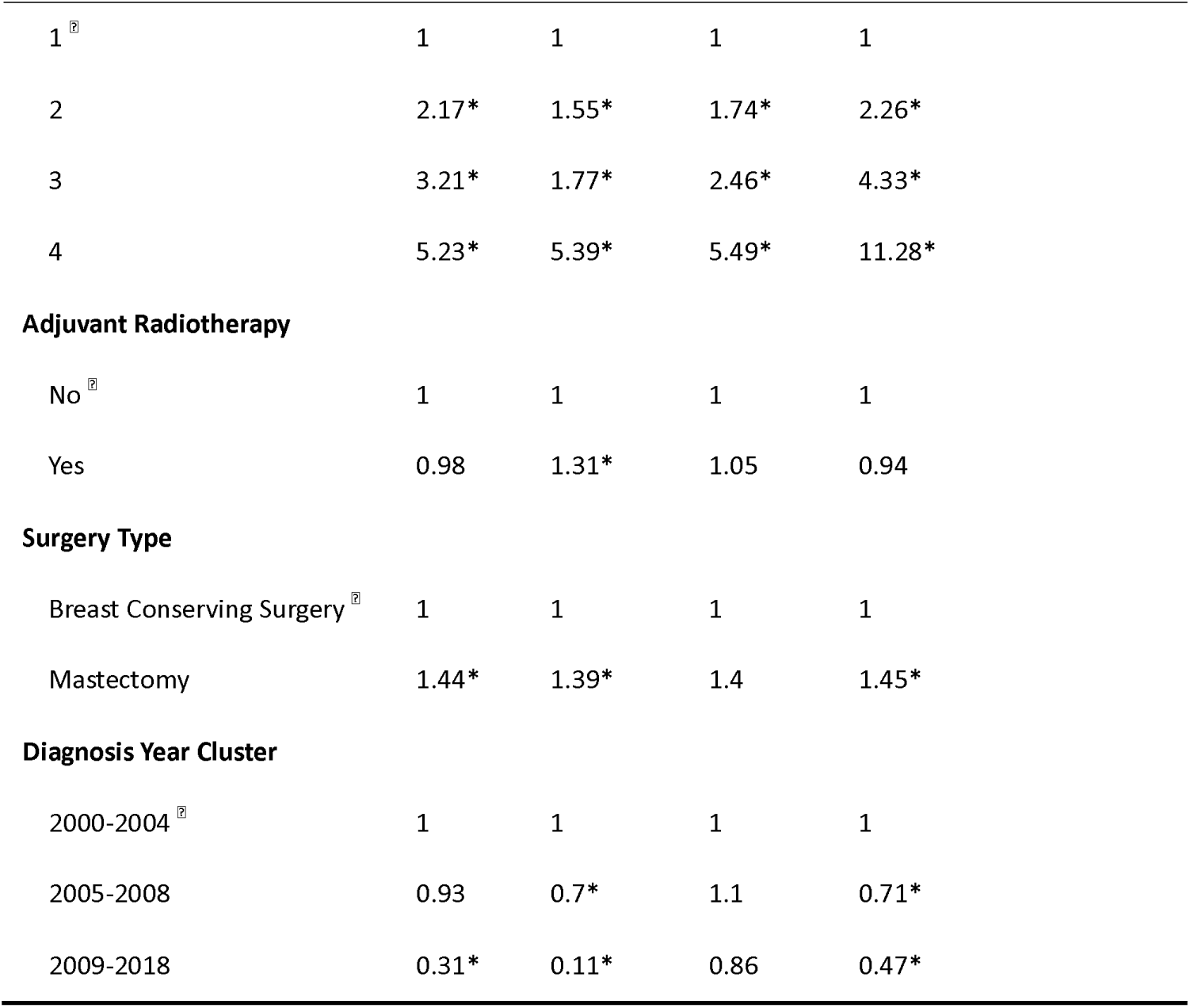
Hazard ratios from multivariable piecewise Cox regression for breast cancer-specific survival. Independent multivariable Cox regression models were fit for sequential segments of follow-up time from diagnosis with breakpoints at 5 and 10-years. An asterisk denotes HRs with a corresponding p-value of < 0.05. The subgroups are ordered with the reference groups first, denoted with a ⍰.

|  | Hazard Ratio (Breast cancer-specific survival) |  |  |  |
| --- | --- | --- | --- | --- |
|  | 0-5 year | 5-10 years | 10+ years | Full dataset |
| **Age at Diagnosis group (years)** |  |  |  |  |
| 45-69 1 | 1 | 1 | 1 | 1 |
| ≤ 44 | 1.24* | 1.12 | 1.19 | 0.97 |
| ≥ 70 | 0.98 | 0.62* | 1.18 | 1.38* |
| **Ethnicity** |  |  |  |  |
| European 1 | 1 | 1 | 1 | 1 |
| Māori | 0.96 | 0.93 | 1.14 | 1.11 |
| Pacific Peoples | 0.84 | 1.14 | 1.12 | 1.05 |
| Asian | 0.6* | 0.76 | 0.66 | 0.64* |
| Other/Unknown | 1.08 | 1.12 | 0.53 | 1.38* |
| **Detection Method** |  |  |  |  |
| Screened 1 | 1 | 1 | 1 | 1 |
| Not Screened | 1.39* | 1.74* | 1.53* | 1.52* |
| **Receptor Subtype** |  |  |  |  |
| ER+/HER2- 1 | 1 | 1 | 1 | 1 |
| ER+/HER2+ | 0.95 | 0.9 | 0.62* | 0.85* |
| ER-/HER2+ | 1.39* | 1.01 | 0.27* | 1.14 |
| Triple Negative | 2.32* | 0.99 | 0.32* | 1.5* |
| **Tumor Grade** |  |  |  |  |
| 1 1 | 1 | 1 | 1 | 1 |
| 2 | 2.72* | 2.47* | 1.6* | 2.44* |
| 3 | 4.97* | 2.68* | 1.11 | 4.02* |
| **Disease Stage (AJCC 7)** |  |  |  |  |
|  | 0-5 year | 5-10 years | 10+ years | Full dataset |
| 1 <sup>§</sup> | 1 | 1 | 1 | 1 |
| 2 | 2.17* | 1.55* | 1.74* | 2.26* |
| 3 | 3.21* | 1.77* | 2.46* | 4.33* |
| 4 | 5.23* | 5.39* | 5.49* | 11.28* |
| **Adjuvant Radiotherapy** |  |  |  |  |
| No <sup>§</sup> | 1 | 1 | 1 | 1 |
| Yes | 0.98 | 1.31* | 1.05 | 0.94 |
| **Surgery Type** |  |  |  |  |
| Breast Conserving Surgery <sup>§</sup> | 1 | 1 | 1 | 1 |
| Mastectomy | 1.44* | 1.39* | 1.4 | 1.45* |
| **Diagnosis Year Cluster** |  |  |  |  |
| 2000-2004 <sup>§</sup> | 1 | 1 | 1 | 1 |
| 2005-2008 | 0.93 | 0.7* | 1.1 | 0.71* |
| 2009-2018 | 0.31* | 0.11* | 0.86 | 0.47* |

By ethnicity, Māori and Pacific women generally had breast cancer survival rates that were comparable to European women, with some variability across different time periods post-diagnosis. The risk of dying of breast cancer for Māori moved from HR 0.93 for those with 5-10 years follow-up to HR 1.14 for those with 10+ years follow-up (not statistically significant). At all time intervals, Asian women had reduced risk compared to European women (overall HR 0.64) (Table ).

Women whose breast cancer was detected after presenting with symptoms had a higher risk of dying from breast cancer than women who were diagnosed by screening, at all time intervals, with an overall HR of 1.52. Women who had a mastectomy had consistently higher mortality risks (HR 1.45 overall) compared to those had breast-conserving surgery as their most advanced surgery (Table 2).

Receptor subtype analysis indicated that women with triple-negative tumors faced the highest risk of death from breast cancer up to 5 years after diagnosis (HR 2.32). However, from 5 to 10 years, their mortality rates were similar to those of women with other subtypes (Table 2). For women surviving beyond 10 years, those with ER+/HER2-tumors exhibited the highest risk of dying from breast cancer. In contrast, those with ER-tumors, either ER-/HER2+ or triple-negative, had the lowest risks (HR 0.27 and 0.32, respectively).

Women with higher tumor grades had a greater risk of dying from breast cancer, especially those with Grade 3 tumors, who exhibited an initial HR of 4.97 (Table 2). However, this risk decreased over time; for those surviving to 10 years, the risk of dying approached that of women diagnosed with Grade 1 tumors (HR 1.11) (Table 2). Compared to women diagnosed with stage 1 disease, all other stages had higher mortality risks at all time intervals, especially those women diagnosed with Stage 4 disease with an HR reaching 11.28 overall (Table 2). Women diagnosed more recently (2009-2018) showed significantly improved BCSS rates (overall HR 0.47) compared to those diagnosed in earlier periods.

Next, piecewise Cox regression analysis of LRR was performed. Even after adjusting for tumor receptor subtype, the multivariate model showed that women diagnosed ≤ 44 years were at greater risk of having a LRR compared to women diagnosed between 45-69 years (overall HR 1.42, Table 3). Their greatest risk of a LRR occurred between 5-10 years after diagnosis (HR 2.29). By ethnicity, Māori had a greater overall risk of a LRR than European women (HR 1.27), with their greatest risk at 10 years after diagnosis (HR 1.26; Table 3).

**Table 3.** Hazard ratios from multivariable piecewise Cox regression for time to locoregional recurrence. Independent multivariable Cox regression models were fit for sequential segments of follow-up time from diagnosis with breakpoints at 5 and 10 years. An asterisk denotes HRs with a corresponding p-value of < 0.05. Region is included as a stratum variable to adjust for region but not generate coefficients. The subgroups are ordered with the reference groups first, denoted with a ⍰.

|  | Hazard Ratio (Locoregional recurrence) |  |  |  |
| --- | --- | --- | --- | --- |
|  | 0-5 year | 5-10 years | 10+ years | Full dataset |
| **Age at Diagnosis group (years)** |  |  |  |  |
| 45-69 □ | 1 | 1 | 1 | 1 |
| $\leq 44$ | 1.54* | 2.29* | 1.8* | 1.42* |
| $\geq 70$ | 0.58* | 0.55* | 0.81 | 1 |
| **Ethnicity** |  |  |  |  |
| European □ | 1 | 1 | 1 | 1 |
| Māori | 1.09 | 0.96 | 1.26 | 1.27* |
| Pacific Peoples | 0.77 | 1.29 | 0.2 | 0.87 |
| Asian | 0.78 | 1.2 | 0.37 | 0.89 |
| Other/Unknown | 0.59* | 0 | 1.89 | 0.79 |
| **Detection Method** |  |  |  |  |
| Screened □ | 1 | 1 | 1 | 1 |
| Not Screened | 1.5* | 0.87 | 1.23 | 1.34* |
|  | 0-5 year | 5-10 years | 10+ years | Full dataset |
| **Receptor Subtype** |  |  |  |  |
| ER+/HER2- <sup>Ⓢ</sup> | 1 | 1 | 1 | 1 |
| ER+/HER2+ | 1.24 | 1 | 0.82 | 1.03 |
| ER-/HER2+ | 1.64* | 0.92 | 0.92 | 1.35* |
| Triple Negative | 2.61* | 1.06 | 0.51 | 1.77* |
| **Tumor Grade** |  |  |  |  |
| 1 <sup>Ⓢ</sup> | 1 | 1 | 1 | 1 |
| 2 | 1.08 | 0.93 | 0.89 | 1.12 |
| 3 | 1.59* | 1.19 | 0.36* | 1.61* |
| **Disease Stage (AJCC 7)** |  |  |  |  |
| 1 <sup>Ⓢ</sup> | 1 | 1 | 1 | 1 |
| 2 | 1.08 | 0.91 | 1.47 | 1.3* |
| 3 | 1.57* | 0.8 | 1.33 | 2.69* |
| **Adjuvant Radiotherapy** |  |  |  |  |
| No <sup>Ⓢ</sup> | 1 | 1 | 1 | 1 |
| Yes | 0.4* | 0.54* | 0.52 | 0.34* |
| **Surgery Type** |  |  |  |  |
| Breast Conserving Surgery <sup>Ⓢ</sup> | 1 | 1 | 1 | 1 |
| Mastectomy | 0.62* | 0.34* | 0.32* | 0.43* |
| **Diagnosis Year Cluster** |  |  |  |  |
| 2000-2004 <sup>Ⓢ</sup> | 1 | 1 | 1 | 1 |
| 2005-2008 | 1.24 | 0.83 | 0.39* | 0.74* |
| 2009-2018 | 0.43* | 0.05* | 0.12* | 0.47* |

**Table 4.** Hazard ratios (HR) from multivariable piecewise Cox regression for distant metastasis-free survival. Independent multivariable Cox regression models were fit for sequential segments of follow-up time from diagnosis with breakpoints at 5 and 10 years. An asterisk denotes hazard ratios with a corresponding p-value of< 0.05.

|  | Hazard Ratio (Distant metastasis-free survival) |  |  |  |
| --- | --- | --- | --- | --- |
|  | 0-5 years | 5-10 years | 10+ years | Full dataset |
| Age at diagnosis ≤ 44 | 1.38* | 1.19 | 1.31 | 1.19* |
| Age at diagnosis ≥ 70 | 0.81* | 0.42* | 1.53* | 1.2* |
| Ethnicity Māori | 0.86* | 0.98 | 1.45 | 1.13* |
| Ethnicity Pacific Peoples | 0.82* | 0.99 | 0.74 | 0.96 |
| Ethnicity Asian | 0.76* | 0.82 | 0.72 | 0.68* |
| Ethnicity Other/Unknown | 1.27* | 1.62 | 1.39 | 1.62* |
| Region Central | 0.88 | 0.75 | 0.35 | 0.9 |
| Region Midland | 0.97 | 0.91 | 0.91 | 1.09* |
| Region Southern | 0.81* | 1.03 | 1.44 | 0.99 |
| Detection method Not Screened | 1.51* | 1.73* | 1.32 | 1.48* |
| Detection method Unknown | 1.02 | 0.96 | 1.94 | 1.31 |
| Grade 2 | 2.29* | 2.02* | 1.87* | 2.44* |
| Grade 3 | 3.83* | 1.96* | 1.27 | 3.87* |
| Receptor subtype ER+/HER2+ | 1.14* | 0.8 | 0.56 | 0.9 |
| Receptor subtype ER-/HER2+ | 1.21* | 0.69 | 0.44 | 1.02 |
| Receptor subtype Triple Negative | 1.65* | 0.8 | 0.27* | 1.21* |
| Receptor subtype Unknown | 1.11 | 0.89 | 0.84 | 0.87* |
| Stage 2 | 2.17* | 1.73* | 1.48* | 2.37* |
| Stage 3 | 3.36* | 2.03* | 2.3* | 4.61* |
| Stage 4 | 20.69* | 3.09* | 3.76* | 27.68* |
| Diagnosis year cluster 2009-2021 | 0.36* | 0.1* | 0.33* | 0.62* |

Having a mastectomy (HR 0.43) or radiation therapy (HR 0.34) were associated with a reduced risk of LRR compared to having breast conserving surgery and not receiving radiotherapy, respectively. Women with ER-breast cancers followed a similar trend in LRR over time as with BCSS, where initially these women had a higher risk of LRR, which then reduced as follow-up time increased. In the LRR case, this trend also held for women with ER+/HER2+ breast cancer receptor subtypes. Tumor grade was inversely associated with late recurrence, more than 10 years after diagnosis, despite adjusting for tumor receptor subtype.

To highlight trends in covariate HRs for each segment as time from diagnosis increased, the risk of LRR progressively increased for women with the ER+/HER2-subtype compared to other receptor subtypes. Conversely, the risk of LRR for women with grade 2 and 3 tumors progressively decreased compared to women with grade 1 tumors. Other covariates such as surgery type (mastectomy) and having adjuvant radiotherapy demonstrated steady risk of LRR as time from diagnosis increased.

There are some notable differences in the pattern of HRs for LRR compared to BCSS. For example, women diagnosed with breast cancer ≤44 years had a higher risk of LRR compared to those diagnosed between 45-69 years (HR 1.41). However, this pattern is reversed when considering BCSS, where women diagnosed ≤44 years had survival rates similar to those aged 45–69 (HR 0.98). In contrast, women diagnosed >69 years had a similar risk of LRR (HR 0.98) but experienced worse survival outcomes (HR 1.37) compared to the 45–69 age group.

Compared to European women (reference group), the risk of LRR for Pacific women (HR 0.87) was similar to Asian women (HR 0.89)), despite having worse BCSS (HR 1.05 for Pacific women and HR 0.64 for Asian women; Table ). In contrast, wāhine Māori had similar BCSS to European women (HR 1.11), but an increased risk of LRR (HR 1.27). Compared to all women with the ER+/HER2-receptor subtype, those with ER+/HER2+ receptor subtype had worse BCSS (HR 0.85) and a similar risk of LRR (HR 1.03), whereas those with ER-/HER2+ tumors had similar survival (HR 1.12) yet higher risk of LRR (HR 1.35). Finally, women who had a mastectomy had a higher risk of BCSS (HR 1.46), despite a lower risk of LRR (HR 0.43) compared to those women who had breast conserving surgery.

### 3.4 Landmark analysis

Based on the results above, we focused on the associations between clinicopathological breast cancer features and patient outcomes and how these changed at various time points after diagnosis. This analysis involved studying only those patients who survived to either 5 or 10 years after diagnosis.

Landmark analysis, focused on breast cancer receptor subtypes and adjusted for age at diagnosis, disease stage, and tumor grade, revealed an interesting change in risk factors over time. Analyzing all women from time of diagnosis showed that those with ER-breast cancer had the worst BCSS and LRR rates, particularly those with triple negative breast cancer (Figures 3A-3B). However, for women that survived to or had no LRR by 5 years, those with ER-tumors had better outcomes than those women with ER+ tumors (Figures 3A-3B). Interestingly, tumor HER2 status did not appear to influence BCSS for women surviving to or LRR-free at 5 years after diagnosis, evidenced by the curves for women with ER+ subtypes and ER-subtypes being very similar, irrespective of tumor HER2 status (Figures 3A-3B). These observations were similar for women that survived to or had no LRR by 10 years after diagnosis. Compared to all other receptor subtypes, women with triple-negative disease who survived to 10 years were less likely to die, in contrast to those who had ER+/HER2-tumors, who exhibited the poorest prognosis (Figure 3C). Among patients who had no LRR by 10 years, the differences in recurrence rates by subtype were minimal (Figure 3D).

**Figure 3.**
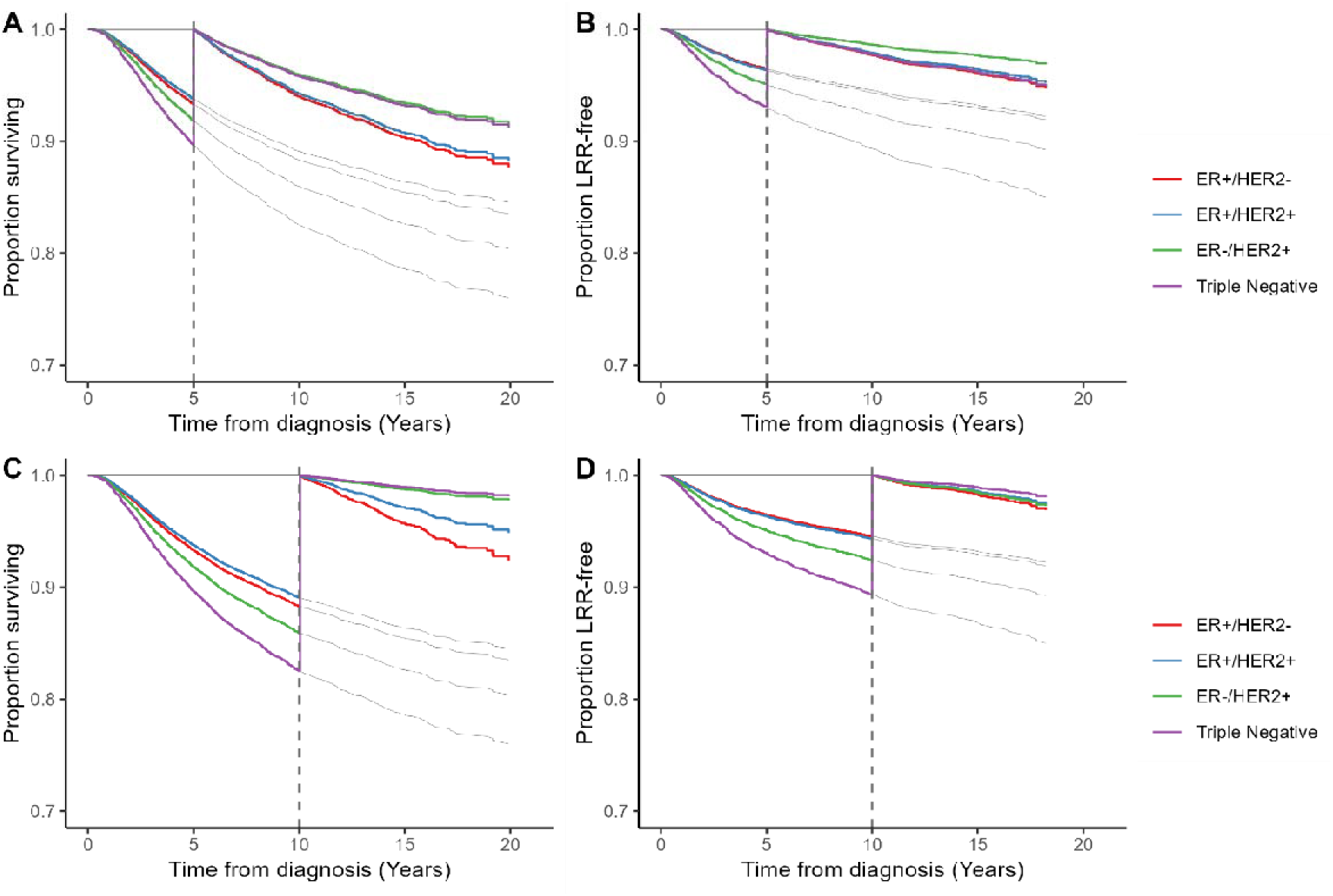
Landmark analysis for breast cancer-specific survival (BCSS) and LRR-free survival across breast tumor receptor subtypes. (A) 5-year landmark with BCSS, (B) 5-year landmark with LRR, (C) 10-year landmark with BCSS, (D) 10-year landmark with LRR. Dashed vertical lines indicate landmark times. Grey lines depict outcomes extending from the time of diagnosis.

### 3.5 Features associated with early and late locoregional recurrence

Logistic regression was used to study the influence of features on early (≤ 5 years) and late LRR ( ≥ 10 years) (Figure ). Women receiving radiation therapy had reduced odds of early LRR compared to those that did not have radiation therapy. Women with stage 3 disease had increased odds of early LRR irrespective of breast cancer receptor subtype. Compared to women having breast conserving surgery, those who had a mastectomy had reduced odds of early LRR for ER+ receptor subtypes. Compared to women diagnosed between 45-69 years, those diagnosed ≤ 44 years had increased odds of both early and late LRR if they had an ER+/HER2-tumor receptor subtype.

Next, given the interest in understanding features that might be associated with late recurrence in women with ER+ tumors, logistic regression was performed, looking LRR < 5 years, ≥ 10 years, or ≥ 15 years. This showed that compared to women diagnosed between 45-69 years, those diagnosed ≤ 44 years had increased odds of having a LRR before 5 years, and ≥ 10 years after diagnosis, but not ≥ 15 years (Figure ). Wāhine Māori had increased odds of early LRR compared to European women, within the ER+ subgroup.

Compared to women not having radiation therapy, those that did have radiation therapy had reduced odds of early LRR, but there was no difference between the groups having LRR ≥ 10 years, suggesting that the benefits of radiation therapy is to minimize early recurrences. However, analysis by surgery type showed that those women having a mastectomy had reduced odds of LRR at all three timepoints assessed compared to women having breast conserving surgery (Figure 5). The most significant feature associated with late LRR in women with ER+ breast tumors was disease stage. Compared to women diagnosed with stage 1 disease, those with stage 2 breast cancer had markedly increased odds of having a LRR ≥ 15 years (OR = 7.89; 95% CI 6.68, 9.09; Figure 5). This was not observed for women with stage 3 disease.

**Figure 4.**
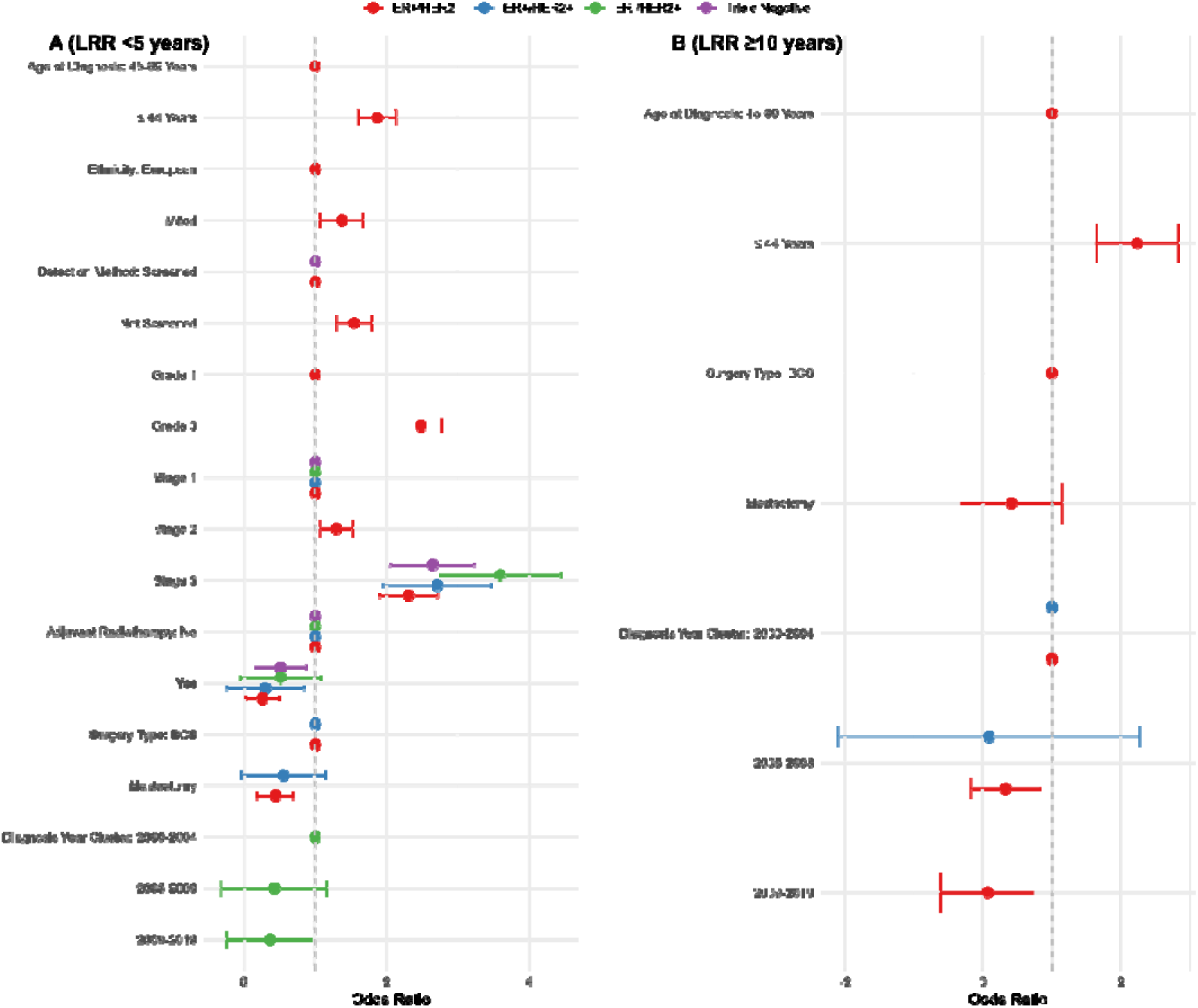
Features associated with early and late locoregional recurrence by tumor receptor subtype. Odds ratios are presented from logistic regression for early LRR within 5 years of diagnosis (A), and late LRR more than 10 years after diagnosis (B). Error bars represent the 95% confidence interval for each estimate. For figure clarity, only statistically significant estimates (p ≤0.05) are presented, i.e. those with p > 0.05 have been removed from the figure. The reference groups for each feature are as follows: Age at Diagnosis = 45-69 Years, Ethnicity = European, Detection Method = Screened, Grade = Grade 1, Stage = Stage 1, Receipt of Adjuvant Radiotherapy = No, Surgery Type = Breast Conserving Surgery (BCS), Diagnosis Year Cluster = 2000-2004.

**Figure 5.**
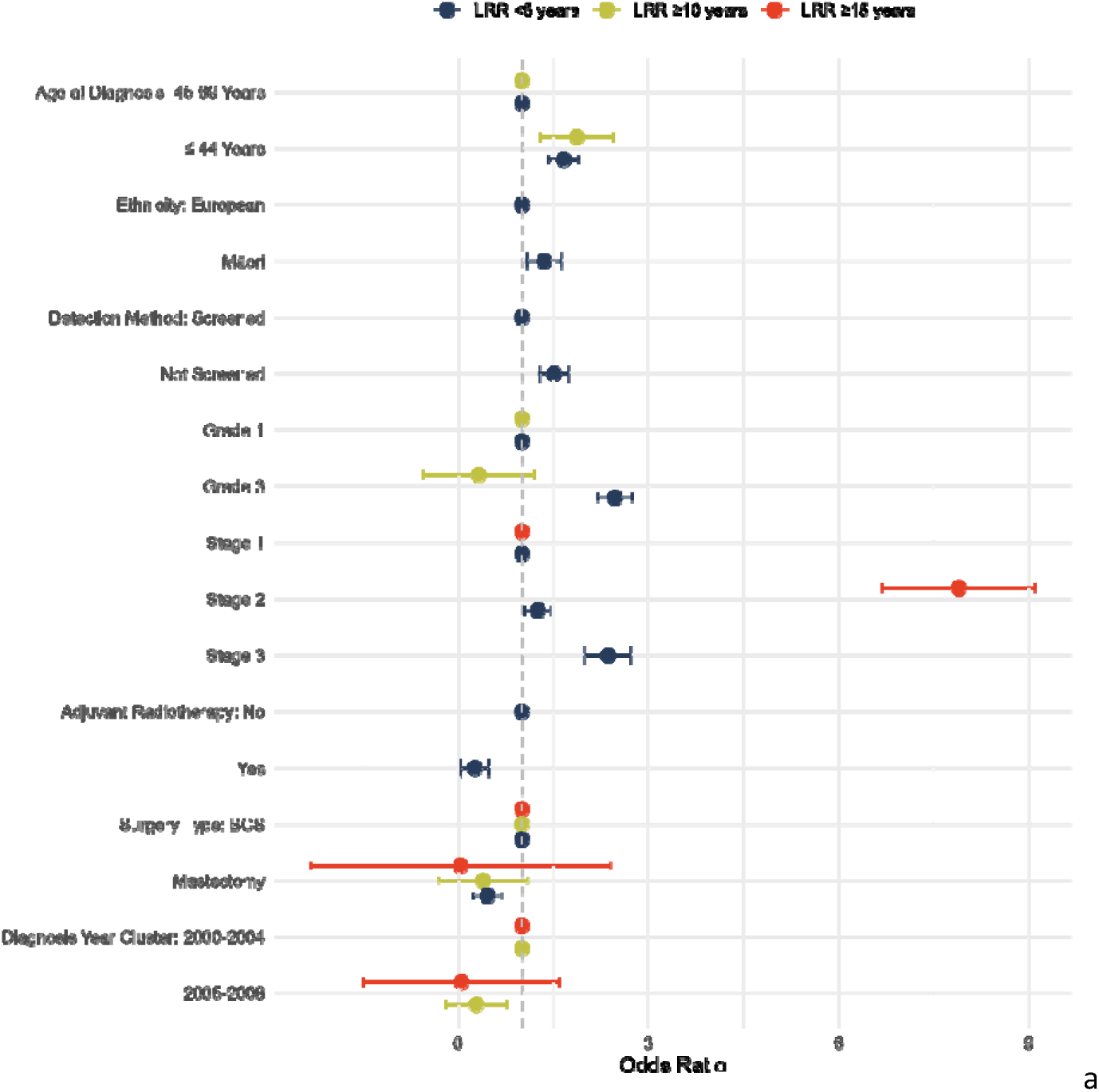
Features associated with early and late locoregional recurrence within ER+ tumor receptor subtypes. Odds ratios are presented from logistic regression for early LRR within 5 years of diagnosis (blue), late LRR ≥ 10 years after diagnosis (yellow), and LRR ≥ 15 years after diagnosis (red). Error bars represent the 95% confidence interval for each estimate. For figure clarity, only statistically significant estimates (p ≤0.05) are presented, i.e. those with p > 0.05 have been removed from the figure. The reference groups for each feature are as follows: Age at Diagnosis 45-69 Years, Ethnicity European, Detection Method Screened, Grade 1, Stage 1, Adjuvant Radiotherapy No, Surgery Type Breast Conserving Surgery (BCS), Diagnosis Year Cluster 2000-2004.

## 4. Discussion

This study revealed how breast cancer prognosis changes over time through a complex interplay of individual risk factors. While the literature is often focused on survival from treatment or diagnosis (Gautier et al., 2022; Lawrenson, Lao, Campbell, Harvey, Seneviratne, et al., 2018), our findings demonstrate a significant risk profile reversal for hormone receptor positive tumors at 5 years post-diagnosis (Figure 3). These results have important implications for long term screening and clinical consultations. Our analysis also uncovered distinct ethnic patterns, with Māori women showing comparable survival but increased LRR risk compared to European women, while Asian women demonstrated consistently better survival outcomes.

Adjusted Kaplan-Meier curves revealed improved prognosis as diagnosis year increased. K-medians survival clustering identified three distinct patient groups, with outcomes progressively improving in more recent diagnosis cohorts. Several factors likely contributed to this improvement, including treatment advances, technological developments, and evolving clinical practices.

The introduction of trastuzumab (Herceptin) in NZ represents one such advance—initially funded for metastatic breast cancer in 2005 with limited earlier access from 2002, followed by full funding for early-stage breast cancer in 2007 (Gabe et al., 2012; Lawrenson, Lao, Campbell, Harvey, Brown, et al., 2018; Manning, 2011; Metcalfe et al., 2007). While significant, trastuzumab’s impact was limited to the approximately 15% of HER2+ patients in our cohort. Additional contributing factors likely include the implementation of Quality Performance Indicators (QPIs) following the Royal Australasian College of Surgeons breast cancer Key Performance Indicators introduction in 2004, and the 2004 extension of BreastScreen Aotearoa’s free screening to women aged 45-69 years (previously 50-64 years).

Landmark analysis demonstrated that women with triple negative breast cancer had an increased risk of breast cancer death and LRR within 5 years compared to women with ER+/HER2-tumors. Similarly, women diagnosed under 45 years, those detected symptomatically, and those with higher tumor grade and disease stage showed increased early mortality risk. These findings align with previous NZ-based cohort analyses (Gautier et al., 2022; Lawrenson, Lao, Campbell, Harvey, Seneviratne, et al., 2018; Seneviratne, Campbell, Scott, Shirley, Peni, et al., 2015) and international studies associating early recurrence with larger tumors, increased nodal involvement, and high grade and stage disease (Murata et al., 2019; Rothschild et al., 2023; Yun et al., 2020).

Notably, women with triple negative disease who survived beyond 5 years demonstrated decreased risk compared to those with ER+/HER2-receptor subtype at both 5- and 10-year landmarks for BCSS. This finding is particularly meaningful as approximately 69% of women with triple negative disease (1,462 of 2,128) were included in the 5-year landmark cohort, indicating this favorable late prognosis applies to a substantial proportion of this population. This risk reversal may reflect treatment influences, as response to neoadjuvant treatment significantly affects survival in triple negative disease, likely due to increased tumor proliferation enhancing chemotherapy efficacy (Liedtke et al., 2023). While the absence of targetable receptors results in poorer initial prognosis for triple negative disease, these patients tend to be younger with fewer comorbidities, potentially enhancing long-term survival.

Our study revealed complex ethnic patterns of BCSS and LRR risk. Particularly noteworthy was the pattern for Māori women, whose hazard ratio for BCSS increased from 0.93 (5-10 years follow-up) to 1.14 (beyond 10 years). This shifting risk profile may partially reflect higher LRR rates (HR 1.27) compared to European women, suggesting that extended follow-up provides sufficient time for recurrences to impact survival outcomes. Asian women demonstrated significantly better BCSS and lower LRR rates compared to European women, while Pacific women showed a more complex pattern—lower LRR rates but slightly worse BCSS—highlighting the need for targeted interventions and further investigation.

Increasing tumor grade was inversely associated with late LRR risk (≥10 years post-diagnosis). Previous research has shown higher grade correlates with reduced relative risk of late recurrence (≥5 years), with relative risks of 0.39 for grade 2 and 0.46 for grade 3 compared to grade 1 cancers (Wangchinda & Ithimakin, 2016). This inverse relationship may result from more aggressive treatment approaches for higher-grade cancers. Although our multivariable model adjusted for radiotherapy and surgery type, it did not account for systemic therapies due to complexities in type, timing, and dosage—a limitation warranting further study.

In women with ER+ tumors, stage 2 disease was associated with significantly greater odds of late recurrence (≥15 years post-diagnosis). This finding may reflect clinical challenges in treating “intermediate risk” patients, with possible undertreatment contributing to increased late recurrence odds.

While younger women (≤44 years) showed greater LRR risk than those diagnosed at 45-69 years, their BCSS rates were similar. This pattern may reflect more aggressive tumor characteristics in younger patients, though our multivariable model controlled for many such factors. Younger women may better tolerate aggressive treatments, contributing to survival outcomes comparable to middle-aged patients. Women receiving mastectomy showed lower recurrence risk—likely due to more complete local resection—but worse BCSS compared to those receiving breast-conserving surgery, potentially highlighting neoadjuvant treatment benefits before breast conservation. Other factors include potential cohort differences, as mastectomy patients often present with more advanced disease and larger tumors.

Landmark analysis provides risk estimations more relevant to specific cohorts rather than including all patients in full models. Its benefits include accounting for covariate non-linearity over time by segmenting follow-up periods and providing more accurate risk assessments that consider how long a patient has been event-free. This approach appropriately excludes patients with poorer outcomes (like incurable disease) that might skew results, though this narrower applicability comes at the cost of reduced statistical (Putter & van Houwelingen, 2017).

When interpreting hazard ratios (Hernán, 2010; Sashegyi & Ferry, 2017; Spruance et al., 2004), which represent instantaneous relative risk assumed constant over time in Cox regression (Loop & Zhang, 2013), context is essential. For example, women with triple negative disease showed a hazard ratio of 1.49 compared to those with ER+/HER2-tumors, suggesting approximately 1.5 times the mortality risk. However, examining absolute risk measures reveals 5-year BCSS of 0.90 (95% CI 0.89, 0.91) for triple negative disease versus 0.93 (95% CI 0.93, 0.94) for ER+/HER2-tumors (Figure 3A). This underscores that hazard ratios alone don’t fully represent clinically relevant risk, and conversion to absolute risk remains essential.

## Limitations

While Te Rēhita provides a comprehensive resource, our analysis has several limitations. The predominantly urban population likely under-represents women from higher deprivation areas, including many wāhine Māori, potentially biasing analysis toward urban Māori women despite evidence that rural Māori women experience worse breast cancer outcomes (Lawrenson et al., 2016). Our models excluded systemic therapies due to complexities in treatment schedules and types, though treatment effects likely explain some observed survival variations. Additional treatment biases include younger women more frequently receiving neoadjuvant therapy, treatment-stage interactions, and personal patient choices. Furthermore, our models omitted potentially valuable information on histological features, comorbidities, symptoms, treatment timing, and private versus public healthcare access—factors future research should consider.

## Ethical Approval

Ethical approval was obtained from the Auckland Health Research Ethics Committee (AHREC) (Ref. AH2800) before the commencement of research.

## Data Availability

Data is available upon reasonable request from the Breast Cancer Foundation National Breast Cancer Registry Clinical Advisory Group.

## Supplementary

**Supplementary Table 1.** Clinicopathological features by diagnosis year cluster, where clusters represent year groups with similar median survival. P values were calculated using a Kruskal-Wallis test for numeric variables (Age at Diagnosis), and a Chi squared test for all remaining categorical variables. Proportion within each cluster (column) is shown in brackets for categorical variables, and standard deviation (SD) within brackets for the continuous variable (age at diagnosis). Women with breast cancer detected following symptoms (as opposed to breast screening) were defined as having detection method “Not Screened”. Breast cancer cases were staged according to the 7th edition of American Joint Committee on Cancer TNM staging criteria.

|  | 2000-2004<br>(n=1,440) | 2005-2008<br>(n=3,204) | 2009-2019<br>(n=16,930) | P-value |
| --- | --- | --- | --- | --- |
| **Age at Diagnosis (years)** |  |  |  |  |
| Mean (SD) | 55.2 (13.5) | 56.9 (13.0) | 58.1 (12.5) | <0.001 |
| Median [Min, Max] | 54.0 [20.0, 97.0] | 56.0 [21.0, 96.0] | 58.0 [20.0, 98.0] |  |
| **Age at Diagnosis group (years)** |  |  |  |  |
| 45-69 | 878 (61.0%) | 2,117 (66.1%) | 11,784 (69.6%) | <0.001 |
| 20-44 | 337 (23.4%) | 549 (17.1%) | 2,154 (12.7%) |  |
| 70-98 | 225 (15.6%) | 538 (16.8%) | 2,992 (17.7%) |  |
| **Ethnicity** |  |  |  |  |
| European | 1,129 (78.4%) | 2,357 (73.6%) | 12,431 (73.4%) | <0.001 |
| Māori | 115 (8.0%) | 327 (10.2%) | 1,652 (9.8%) |  |
| Pacific Peoples | 58 (4.0%) | 198 (6.2%) | 1,035 (6.1%) |  |
| Asian | 100 (6.9%) | 243 (7.6%) | 1,540 (9.1%) |  |
| Other/Unknown | 38 (2.6%) | 79 (2.5%) | 272 (1.6%) |  |
| **Detection Method** |  |  |  |  |
| Screened | 459 (31.9%) | 1,230 (38.4%) | 8,038 (47.5%) | <0.001 |
| Not Screened | 981 (68.1%) | 1,974 (61.6%) | 8,892 (52.5%) |  |
| **Region (domicile at diagnosis)** |  |  |  |  |
| Northern | 1,099 (76.3%) | 2,469 (77.1%) | 8,671 (51.2%) | <0.001 |
| Central | 0 (0%) | 0 (0%) | 2,780 (16.4%) |  |
| Midland | 341 (23.7%) | 735 (22.9%) | 2,443 (14.4%) |  |
| Southern | 0 (0%) | 0 (0%) | 3,036 (17.9%) |  |
| **Receptor Subtype** |  |  |  |  |
| ER+/HER2- | 853 (59.2%) | 2,261 (70.6%) | 13,063 (77.2%) | <0.001 |
| ER+/HER2+ | 187 (13.0%) | 298 (9.3%) | 1,697 (10.0%) |  |
| ER-/HER2+ | 139 (9.7%) | 231 (7.2%) | 717 (4.2%) |  |
| Triple Negative | 261 (18.1%) | 414 (12.9%) | 1,453 (8.6%) |  |
| **Tumour Grade** |  |  |  |  |
| 1 | 197 (13.7%) | 772 (24.1%) | 3,841 (22.7%) | <0.001 |
| 2 | 688 (47.8%) | 1,452 (45.3%) | 8,077 (47.7%) |  |
| 3 | 555 (38.5%) | 980 (30.6%) | 5,012 (29.6%) |  |
| **Disease Stage (AJCC 7)** |  |  |  |  |
| 1 | 730 (50.7%) | 1,763 (55.0%) | 9,877 (58.3%) | <0.001 |
| 2 | 569 (39.5%) | 1,127 (35.2%) | 5,640 (33.3%) |  |
| 3 | 85 (5.9%) | 188 (5.9%) | 985 (5.8%) |  |
| 4 | 56 (3.9%) | 126 (3.9%) | 428 (2.5%) |  |
| **Adjuvant Radiotherapy** |  |  |  |  |
| No | 499 (34.7%) | 1,130 (35.3%) | 5,875 (34.7%) | 0.822 |
| Yes | 941 (65.3%) | 2,074 (64.7%) | 11,055 (65.3%) |  |
| **Surgery Type** |  |  |  |  |
| Breast Conserving Surgery | 636 (44.2%) | 1,512 (47.2%) | 8,759 (51.7%) | <0.001 |
| Mastectomy | 804 (55.8%) | 1,692 (52.8%) | 8,171 (48.3%) |  |

**Supplementary 1:**
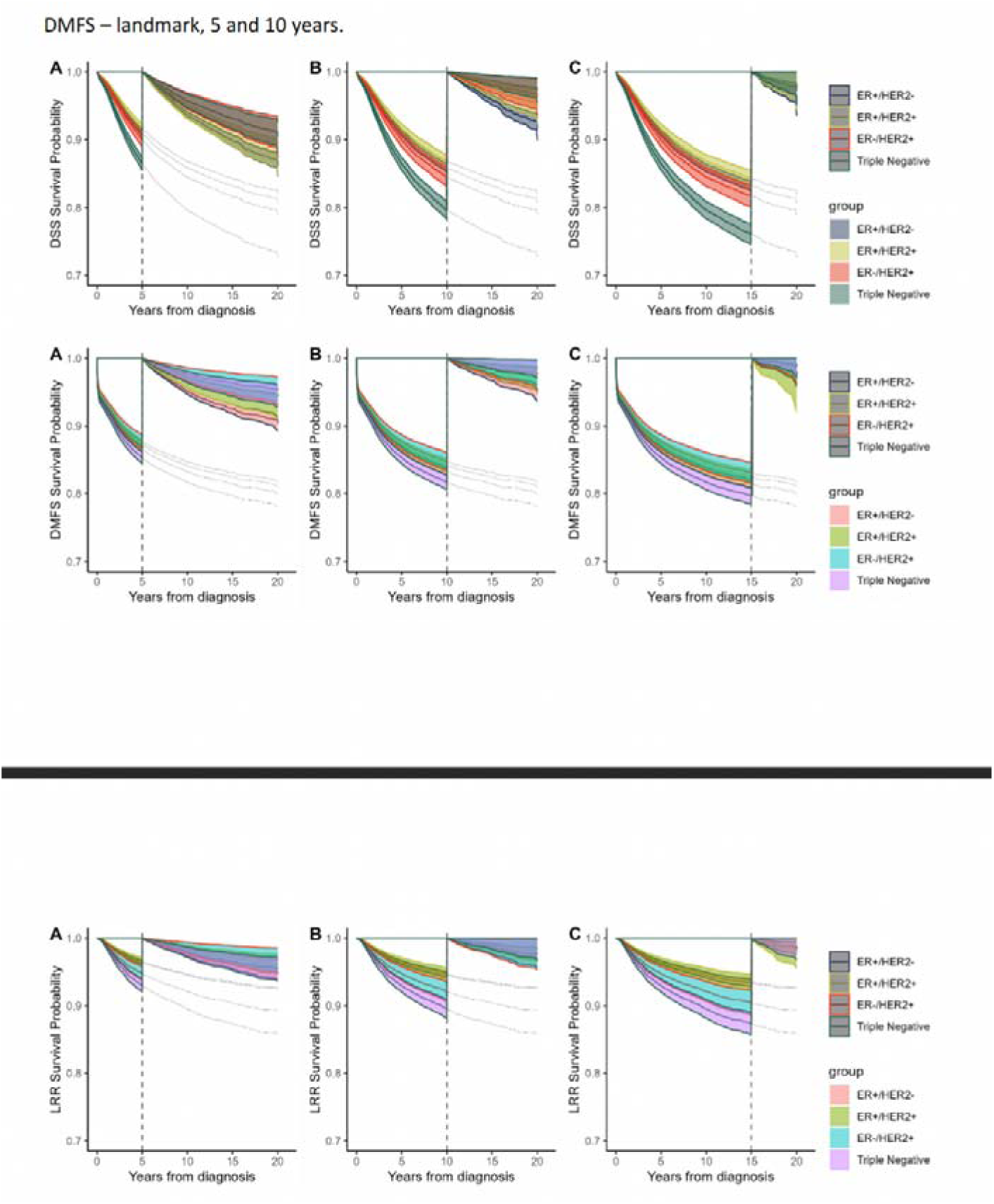
For reference - Figure 2 with confidence intervals.

